# Are older adults using ChatGPT for medical advice? Results from a cross-sectional survey study

**DOI:** 10.64898/2026.01.01.25342468

**Authors:** Meghan Reading Turchioe, Afra Shamnath, Jarrott Mayfield, Orson Xu, David Slotwiner, Natalie Benda

## Abstract

General-purpose large language models (LLMs) like ChatGPT are increasingly used for medical advice despite lacking medical training and frequently producing incorrect or unsafe output. Older adults’ health information-seeking behaviors using LLMs remain poorly characterized. We conducted a cross-sectional survey of 574 US adults aged 50+ recruited via Prolific, balanced by sex and race. Participants reported health information sources, ChatGPT and PubMed use, demographics, and health literacy. Most participants (92%) searched online for health information. All had heard of ChatGPT, and 63% used it for medical information, compared to 44% who had heard of PubMed and 39% who used it. Those with inadequate health literacy had higher odds of ChatGPT use for medical advice (AOR 2.36, 95% CI 1.30-4.52) versus those with adequate health literacy. In conclusion, with more than half of older adults using LLMs for medical advice, the development of safer, purpose-trained medical LLMs is warranted.

## Introduction

Health information-seeking behaviors, referring to ways of gathering health-related information, are associated with the ability to self-manage health and, ultimately, health outcomes.^1^ Adults in the U.S. may use general-purpose large language models (LLMs), such as ChatGPT, for medical advice.^2^ LLMs provide natural language responses to user queries that are easy to understand, but are not trained or tested for medical purposes. One review found 88% of included studies evaluating LLMs for medical purposes reported incorrect or incomplete output, and nearly half reported unsafe output.^3^ Chronic conditions affect 80-90% of adults age 35 and older,^4^ creating substantial medical information needs. However, older adults’ health information-seeking behaviors in the new era of broadly accessible LLMs have not been well characterized. This study investigated health information-seeking behaviors, including LLM use, among US adults aged 50 and older.

## Methods

This cross-sectional survey study used Prolific to recruit participants balanced by sex and reflecting 2020 US Census racial distribution (58% White, 14% Black, 6% Asian, 20% other, and 2% mixed). Eligible participants could read English and were aged 50 and older.

Survey items were adapted from the Health Information National Trends Survey and a separate survey assessing online health information search habits and sources.^5^ We asked about familiarity with and frequency of using ChatGPT (selected because it is the most commonly used LLM with over 700 million users as of September 2025). To contrast this, we also asked about their use of PubMed (a federally supported biomedical library). Participants self-reported demographics and health literacy, completing surveys via Qualtrics. Quality was audited through attention checks and response times. Participants completing the questionnaire and passing attention checks received $16 per hour. The Columbia University Institutional Review Board approved this study.

We used descriptive statistics and logistic regression to examine demographic effects on ChatGPT use for health information (binarized as never versus any use). Analyses were performed using R version 4.1.2.

## Results

Of 622 participants recruited, 48 were excluded (33 age ineligible, 9 incomplete surveys, 6 failed attention checks; 0 excluded for low response times), yielding 574 participants. Mean age was 58 (SD 6.8) years; 51% were female, 70% White, 16% Black or African American, and 10% Hispanic or Latino. Education levels were 30% high school or less, 38% Bachelor’s degree, and 32% graduate degree; 71% had adequate health literacy. Most (93%) used the Internet more than once daily.

Most participants (92%) used the Internet to search for health information (**Table 1**). All but two had heard of ChatGPT (100%), and 63% used it at least once for medical information. Fewer had heard of PubMed (44%) or used it (39%). Other common sources included licensed healthcare professionals (93%) and medical websites (75%). Fewer used government health agencies (40%), news websites (16%), and social media (15%).

**Table 1.** Health information seeking among survey participants (n=574)

|  |  |
| --- | --- |
| **Table 1. Health information seeking among survey participants (n=574)** |  |
| **Tried looking for information about your health from any source** | 553 (96.3%) |
| **Used the Internet for these health-related needs in the last 12 months** |  |
| Look for health or medical information | 525 (91.5%) |
| View medical test results | 349 (60.8%) |
| Made an appointment with a health care provider | 331 (57.7%) |
| Send a message to a healthcare provider | 325 (56.6%) |
| **Heard of ChatGPT** | 572 (100%) |
| **If yes, frequency of ChatGPT use for medical information** |  |
| Often | 99 (17.2%) |
| Sometimes | 154 (26.8%) |
| Once or twice | 111 (19.3%) |
| Never | 208 (36.%) |
| N/A- have not used ChatGPT | 2 (0.3%) |
| **Heard of the online biomedical library, PubMed** | 254 (44.3%) |
| **If yes, frequency of PubMed use for medical information** |  |
| Often | 27 (4.7%) |
| Sometimes | 116 (20.2%) |
| Once or twice | 81 (14.1%) |
| Never | 30 (5.2%) |
| N/A- have not used PubMed | 320 (55.7%) |
| **Sources relied upon for information about personal health** |  |
| A licensed healthcare professional (i.e., doctor or nurse) | 531 (92.5%) |
| Medical website (i.e., WebMD) | 428 (74.6%) |
| Scientific researchers | 336 (58.5%) |
| Government health agencies | 232 (40.4%) |
| Other people with the same health condition | 193 (33.6%) |
| Family, friend, or community member | 177 (30.8%) |
| News website | 93 (16.2%) |
| Social media (i.e., Facebook or Instagram) | 87 (15.2%) |
| Religious organizations and leaders | 15 (2.6%) |

In adjusted models (**Table 2**), Asian participants had lower odds of using ChatGPT for medical information versus White participants (adjusted odds ratio [AOR] 0.36, 95% confidence interval [CI] 0.16-0.78). Participants with graduate degrees (AOR 2.57, 95% CI 1.51-4.43) and Bachelor’s degrees (AOR 1.66, 95% CI 1.01-2.72) had higher odds of using ChatGPT versus those with high school education or less. Those with inadequate health literacy had higher odds of ChatGPT use (AOR 2.36, 95% CI 1.30-4.52) versus those with adequate health literacy.

**Table 2.** Differences in frequency of ChatGPT use for medical advice by sociodemographic characteristics, n=574.

|  |  |  |  |  |  |  |
| --- | --- | --- | --- | --- | --- | --- |
|  | Never (n=208) | Any use (n=364) | Unadjusted OR (95% CI) | p value | Adjusted OR (95% CI) | p value |
| **Age** |  |  |  |  |  |  |
| Reference: 50-59 | 122 (33%) | 246 (67%) | — | — | — | — |
| 60–69 | 67 (41%) | 95 (59%) | 0.70 (0.48–1.03) | 0.070 | 0.76 (0.48–1.21) | 0.247 |
| 70+ | 19 (45%) | 23 (55%) | 0.60 (0.32–1.15) | 0.121 | 0.76 (0.36–1.63) | 0.476 |
| **Sex** |  |  |  |  |  |  |
| Reference: Female | 113 (39%) | 179 (61%) | — | — | — | — |
| Male | 95 (34%) | 185 (66%) | 1.23 (0.87–1.73) | 0.236 | 1.04 (0.69–1.57) | 0.845 |
| **Race** |  |  |  |  |  |  |
| Reference: White | 148 (37%) | 254 (63%) | — | — | — | — |
| Black or African American | 19 (21%) | 72 (79%) | 2.21 (1.30–3.90) | 0.004 | 1.83 (0.98–3.55) | 0.063 |
| Asian | 20 (53%) | 18 (47%) | 0.52 (0.27–1.02) | 0.058 | 0.36 (0.16–0.78) | 0.010 |
| American Indian or Alaska Native | 7 (47%) | 8 (53%) | 0.67 (0.23–1.93) | 0.441 | 0.53 (0.15–1.83) | 0.316 |
| Native Hawaiian or Other Pacific Islander* | 0 (0%) | 1 (100%) | — | — | — | — |
| Prefer not to answer* | 14 (56%) | 11 (44%) | — | — | — | — |
| **Ethnicity** |  |  |  |  |  |  |
| Reference: not Hispanic/Latino | 191 (38%) | 314 (62%) | — | — | — | — |
| Hispanic/Latino | 16 (29%) | 40 (71%) | 1.52 (0.84–2.86) | 0.176 | 1.67 (0.78–3.86) | 0.206 |
| Prefer not to answer* | 1 (9%) | 10 (91%) | — | — | — | — |
| **Education** |  |  |  |  |  |  |
| Reference: High school or less | 85 (50%) | 86 (50%) | — | — | — | — |
| Bachelor's degree | 78 (36%) | 139 (64%) | 1.76 (1.17–2.65) | 0.007 | 1.66 (1.01–2.72) | 0.045 |
| Graduate degree (Master's or higher) | 44 (24%) | 139 (76%) | 3.12 (2.00–4.94) | <0.001 | 2.57 (1.51–4.43) | <0.001 |
| Prefer not to answer* | 1 (100%) | 0 (0%) | — | — | — | — |
| **Finances** |  |  |  |  |  |  |
| Reference: More than enough | 22 (39%) | 35 (61%) | — | — | — | — |
| Enough | 95 (28%) | 245 (72%) | 1.62 (0.89–2.89) | 0.105 | 1.78 (0.92–3.43) | 0.086 |
| Not enough | 83 (52%) | 77 (48%) | 0.58 (0.31–1.07) | 0.087 | 0.65 (0.31–1.34) | 0.244 |
| Prefer not to answer* | 8 (53%) | 7 (47%) | — | — | — | — |
| **Health Literacy** |  |  |  |  |  |  |
| Reference: Adequate | 185 (40%) | 275 (60%) | — | — | — | — |
| Inadequate | 23 (21%) | 89 (79%) | 3.28 (1.93–5.89) | <0.001 | 2.36 (1.30–4.52) | 0.007 |
| **Health insurance coverage in the last year** |  |  |  |  |  |  |
| Reference: Yes, for full year | 185 (39%) | 294 (61%) | — | — | — | — |
| No/ Part of the year | 21 (24%) | 65 (76%) | 1.95 (1.17–3.36) | 0.013 | 2.03 (0.93–4.87) | 0.091 |
| Prefer not to answer* | 2 (29%) | 5 (71%) | — | — | — | — |
| **Type of health insurance of in the past year** |  |  |  |  |  |  |
| Reference: Medicaid | 58 (34.3%) | 111 (65.6%) | — | — | — | — |
| Other than Medicaid | 150 (37.2%) | 253 (62.8%) | 0.87 (0.59–1.27) | 0.483 | 0.73 (0.46–1.17) | 0.198 |

\*Models did not converge due to too few observations.

## Discussion

Over 60% of adults aged 50 and older have used ChatGPT for medical information, with use varying by race, education, and health literacy. By contrast, our recent Prolific study of younger participants (mean age 39) reported lower rates (33%) of ChatGPT use for medical information.^6^ Findings align with our prior work showing that adults with lower healthy literacy are more open to using AI-based tools in their care.^7^ A limitation of this study includes that we recruited participants via an online sample, who may have been predisposed to using AI-based tools and may differ from other adults aged 50 and older. Future work should develop purpose-trained medical LLMs that prioritize safety and usability for older adults with low health literacy.

## Data Availability

Data, statistical code, and the informed consent document will be made available upon reasonable request after completing a data use agreement.

## Author contributions

**Turchioe:** Conceptualization, investigation, writing - original draft, supervision, funding acquisition; **Shamnath:** Formal analysis, writing - review and editing; **Mayfield:** Formal analysis, writing - review and editing; **Xu, Slotwiner**, and **Benda**: Methodology; writing - review and editing.

## Human ethics and consent to participate declarations

The Columbia University Institutional Review Board approved this study (IRB AAAU2028). Participants read an information sheet and consented to participate in this specific study before completing the survey.

---

STROBE Statement–Checklist of items that should be included in reports of ***cross-sectional studies***

STROBE Statement—Checklist of items that should be included in reports of *cross-sectional studies*
|  | Item No | Recommendation | Page No |
| --- | --- | --- | --- |
| Title and abstract | 1 | (a) Indicate the study's design with a commonly used term in the title or the abstract | 1 |
|  |  | (b) Provide in the abstract an informative and balanced summary of what was done and what was found | N/A due to article type |
| Introduction |  |  |  |
| Background/rationale | 2 | Explain the scientific background and rationale for the investigation being reported | 2 |
| Objectives | 3 | State specific objectives, including any prespecified hypotheses | 2 |
| Methods |  |  |  |
| Study design | 4 | Present key elements of study design early in the paper | 2 |
| Setting | 5 | Describe the setting, locations, and relevant dates, including periods of recruitment, exposure, follow-up, and data collection | 2 |
| Participants | 6 | (a) Give the eligibility criteria, and the sources and methods of selection of participants | 2 |
| Variables | 7 | Clearly define all outcomes, exposures, predictors, potential confounders, and effect modifiers. Give diagnostic criteria, if applicable | 2 |
| Data sources/measurement | 8* | For each variable of interest, give sources of data and details of methods of assessment (measurement). Describe comparability of assessment methods if there is more than one group | 2 |
| Bias | 9 | Describe any efforts to address potential sources of bias | 2-3 |
| Study size | 10 | Explain how the study size was arrived at | 3 |
| Quantitative variables | 11 | Explain how quantitative variables were handled in the analyses. If applicable, describe which groupings were chosen and why | 3 |
| Statistical methods | 12 | (a) Describe all statistical methods, including those used to control for confounding | 3 |
|  |  | (b) Describe any methods used to examine subgroups and interactions | N/A |
|  |  | (c) Explain how missing data were addressed | 2-3 |
|  |  | (d) If applicable, describe analytical methods taking account of sampling strategy | N/A |
|  |  | (e) Describe any sensitivity analyses | N/A |
| Results |  |  |  |
| Participants | 13* | (a) Report numbers of individuals at each stage of study—eg numbers potentially eligible, examined for eligibility, confirmed eligible, included in the study, completing follow-up, and analysed | 3 |
|  |  | (b) Give reasons for non-participation at each stage | 3 |
|  |  | (c) Consider use of a flow diagram | N/A due to space |
| Descriptive data | 14* | (a) Give characteristics of study participants (eg demographic, clinical, social) and information on exposures and potential confounders | Table 1 |
|  |  | (b) Indicate number of participants with missing data for each variable of interest | Table 1 |
| Outcome data | 15* | Report numbers of outcome events or summary measures | Table 1 |
| Main results | 16 | (a) Give unadjusted estimates and, if applicable, confounder-adjusted estimates and their precision (eg, 95% confidence interval). Make clear which confounders were adjusted for and why they were included | 3-4 |
|  |  | (b) Report category boundaries when continuous variables were categorized | N/A |
|  |  | (c) If relevant, consider translating estimates of relative risk into absolute risk for a meaningful time period | N/A |
| Other analyses | 17 | Report other analyses done—eg analyses of subgroups and interactions, and sensitivity analyses | N/A |
| **Discussion** |  |  |  |
| Key results | 18 | Summarise key results with reference to study objectives | 4 |
| Limitations | 19 | Discuss limitations of the study, taking into account sources of potential bias or imprecision. Discuss both direction and magnitude of any potential bias | 4 |
| Interpretation | 20 | Give a cautious overall interpretation of results considering objectives, limitations, multiplicity of analyses, results from similar studies, and other relevant evidence | 4 |
| Generalisability | 21 | Discuss the generalisability (external validity) of the study results | 4 |
| **Other information** |  |  |  |
| Funding | 22 | Give the source of funding and the role of the funders for the present study and, if applicable, for the original study on which the present article is based | 5 |

\*Give information separately for exposed and unexposed groups.
**Note:** An Explanation and Elaboration article discusses each checklist item and gives methodological background and published examples of transparent reporting. The STROBE checklist is best used in conjunction with this article (freely available on the Web sites of PLoS Medicine at http://www.plosmedicine.org/, Annals of Internal Medicine at http://www.annals.org/, and Epidemiology at http://www.epidem.com/). Information on the STROBE Initiative is available at www.strobe-statement.org.

